# Neutrophil gelatinase-associated lipocalin (NGAL) is a poor diagnostic marker for sepsis in the ICU – an observational multicentre study

**DOI:** 10.64898/2026.02.12.26346132

**Authors:** Lisa Boström, Sofie Hagström, Jonas Engström, Anders Olof Larsson, Hans Friberg, Maria Lengquist, Attila Frigyesi

## Abstract

**Background:** Sepsis is a major public health challenge, and reliable biomarkers are essential for distinguishing sepsis from other conditions. Neutrophil Gelatinase-Associated Lipocalin (Neutrophil gelatinase-associated lipocalin (NGAL)) has shown promise as a diagnostic marker due to its role in the immune response. This study evaluates plasma NGAL as a diagnostic tool at the time of ICU admission.

**Methods:** We analysed plasma NGAL and C-reactive protein (CRP) levels in 4732 adult patients admitted to four ICUs between 2015 and 2018. All patients were retrospectively screened for Sepsis-3 criteria at ICU admission. The discriminative performance of NGAL and CRP for sepsis was assessed using receiver operating characteristic (ROC) analysis, with NGAL levels adjusted for Chronic kidney disease (CKD) and age. Patients were stratified by renal function.

**Results:** Plasma NGAL levels were significantly higher in septic patients (p<0.001). For the whole cohort, NGAL alone yielded an Area under the curve (AUC) of 0.67 (Confidence interval (CI) 0.66–0.69), CRP yielded an AUC of 0.72 (CI 0.71–0.73, p<0.001), and combining NGAL with CRP nominally improved discriminative performance (AUC 0.74 vs 0.72, p<0.001). Stratified analyses indicated that NGAL, together with CRP, significantly outperformed CRP alone in patients with no kidney injury and those with Acute Kidney Injury (AKI) only. In contrast, differences were not significant in patients with CKD only or CKD and AKI.

**Conclusion:** In this large cohort, NGAL showed modest discrimination for sepsis, with a nominal improvement when combined with CRP. These findings do not indicate that NGAL meaningfully improves sepsis diagnosis in the ICU.

## Introduction

Sepsis remains a significant public health challenge, and improved diagnostic tools are needed [1, 2]. Neutrophil Gelatinase-Associated Lipocalin (Neutrophil Gelatinase-Associated Lipocalin (NGAL)), also known as lipocalin-2, is an intriguing biomarker candidate for diagnosing sepsis. This 198-amino-acid glycoprotein belongs to the lipocalin family and is primarily secreted by immune cells, including neutrophils, macrophages, and dendritic cells [3, 4]. NGAL exists in three forms: a 25 kDa monomer, a 45 kDa dimer, and a 135 kDa heterodimer covalently bound to gelatinase [3]. It is widely distributed across various tissues, such as the kidney, heart, lung, bone marrow, liver, and adipose tissue, and is upregulated in response to inflammatory and metabolic disorders [4, 5].

The monomeric form is the predominant form secreted by renal tubular epithelial cells, and the dimeric form is the predominant form secreted by neutrophils [6]. The heterodimeric form, bound to matrix metalloproteinase-9 (MMP-9), plays a role in extracellular matrix degradation [7].

NGAL’s potential as an early biomarker for Acute Kidney Injury (AKI) has been extensively documented [4, 8–10]. Plasma NGAL levels rise rapidly within two hours of AKI onset, peaking at six hours, offering an earlier indication of kidney injury compared to creatinine [4, 11, 12]. Since 2023, the FDA has approved NGAL as a biomarker for predicting severe AKI in pediatric patients [10].

Beyond its role in kidney injury, NGAL is pivotal in the body’s defence against bacterial infection by sequestering iron-loaded siderophores, thereby inhibiting bacterial growth [7, 13]. This iron-withholding mechanism is crucial to the innate immune response, with NGAL regulating iron-dependent genes [9, 14].

The role of NGAL in sepsis-associated AKI has been explored in several studies, highlighting that NGAL levels are elevated in sepsis [15–18]. Notably, research by Mårtensson et al. and Paul et al. indicate that the association between plasma NGAL and sepsis are independent of AKI [19, 20]. Despite evidence linking NGAL to inflammation and kidney failure [4, 16], studies investigating NGAL as a diagnostic marker for sepsis remain limited, constrained by small sample sizes and diverse clinical settings [19, 21–25].

This study aims to evaluate, within a large cohort of critically ill patients, the utility of NGAL as a clinical biomarker for distinguishing sepsis from non-sepsis. We hypothesize that NGAL serves as a diagnostic marker for sepsis in the Intensive Care Unit (ICU). For comparative analysis, we also examine the well-established sepsis marker, C-reactive Protein (CRP) [26, 27].

## Materials and methods

### Study design and setting

This retrospective observational study used prospectively collected blood samples from the Swecrit biobank [28]. The biobank comprises blood samples from all patients admitted to the ICUs at four hospitals in southern Sweden between 30/03/2015 and 31/12/2018: Skåne University Hospital in Lund and Malmö, Helsingborg Hospital, and Kristianstad Hospital. For the present study, we included blood samples from patients who met the predefined inclusion criteria.

Data were accessed for research purposes in 15/05/2024. The authors had access to pseudonymized data during data analysis.

Ethical approval was obtained from the Ethical Committee at Lund University, Lund, Sweden (EPN 2015/267 and 2017/802), before the study was conducted. The study adhered to all relevant local guidelines and regulations and was conducted in accordance with the Declaration of Helsinki.

The Standards for Reporting Diagnostic Accuracy Studies (STARD) guidelines [29, 30] were followed during the study.

### Participants

This study included all ICU patients ≥ 18 years of age who had a stay >24 hours or died within the first 24 hours and had eligible blood samples in the biobank.

Sepsis was defined as a Sequential Organ Failure Assessment (SOFA) score ≥ 2 within ±1 hour of ICU admission in combination with a retrospective classification of infection according to the Sepsis-3 criteria [31]. Criteria for infection were either 1) culture positivity within ±48 hours of ICU admission, or 2) culture negativity and suspected infection (blood culture sampling within 24 hours of ICU admission with concomitant antibiotic administration) and probable infection according to the Linder-Mellhammar Criteria of Infection (LMCI) [32, 33]. Although some infection-related data were obtained within 24 hours after admission, these variables most likely reflect an infectious process already present at ICU entry. Given that infection rarely progresses from absence to fulminant sepsis within such a short timeframe, this strategy preserves diagnostic relevance.

### Data sources and variables

Clinical data were entered by the treating physician into the PasIva software (used to collect data for the Swedish Intensive Care Registry). These variables include vital signs and other variables needed to calculate the Simplified Acute Physiology Score 3 (SAPS-3) and SOFA scores [34, 35]. Laboratory values, microbiological testing and results were automatically extracted using the hospital’s electronic laboratory system. Medical records were reviewed for information on the administration of antibiotics and LMCI criteria in culture-negative patients. For biomarkers that were measured multiple times, the values closest to ICU admission (within 24 hours) were selected. CRP was used for comparison based on its current role as the most commonly used marker for sepsis [26, 27].

Shock was defined as a cardiovascular SOFA score of 3 or more at ICU admission, equal to using a vasopressor (norepinephrine or epinephrine), combined with a lactate level of >2 mmol/L.

Neutrophil count was analysed based on clinical indication and was not routinely analysed in all patients. Neutropenia was defined as a neutrophil count of < 0.5 × 10^9^. Leukopenia was defined as a White Blood Cell count (WBC) < 3.5 × 10^9^. Body temperature was obtained from the SAPS-3 score and was the highest recorded within ±1 h of ICU admission. Hypothermia was defined as a body temperature <36.0 °C.

The Glomerular Filtration Rate (GFR) was estimated using the Chronic Kidney Disease Epidemiology Collaboration Equation (CKD-EPI) [36].

Chronic kidney disease (CKD) was defined as a pre-admission Estimated Glomerular Filtration Rate (eGFR) <60 mL/min/1.73 m^2^, calculated from the most recent creatinine measurement obtained 7–365 days before ICU admission. NGAL models were adjusted for CKD as a binary covariate, given that CKD may elevate NGAL independently of sepsis. Creatinine measurement obtained 7–365 days before ICU admission was missing in 38% of the patients. In these cases, we assumed the absence of chronic kidney disease.

Acute worsening of kidney function in patients with CKD was defined as an increase in creatinine at admission of more than or equal to 1.5 times baseline creatinine.

Acute kidney injury in patients without CKD was defined as eGFR <60 at admission to the ICU.

Microbiological cultures were defined as clinically relevant if there was growth or detection of a pathogen from a culture taken within ±48 hours of ICU admission, regardless of the anatomical site of culturing. The following culture results were considered clinically irrelevant [37]:

- Yeast fungi from non-sterile anatomical sites (e.g. airways, lower urinary tract, skin lesions)
- Potentially colonising bacteria of the upper or lower respiratory tract, if found in only one airway culture: Moraxella sp., coagulase-negative Staphylococci, viridans (alpha hemolytic) Streptococci
- Potential skin contaminants found in only one blood culture: coagulase-negative Staphylo-cocci, viridans (alpha hemolytic) Streptococci, micrococcus sp., Propionibacterium Acnes, Corynebacterium sp., Bacillus sp.
- Unspecific culture results (e.g. “gram-positive mixed flora”, “vaginal flora”, “skin flora”, “anaerobic mixed flora”) from non-sterile anatomical sites
- Bacterial growth of Clostridium difficile, without detection of toxin
- Pneumococcus antigen tests from urinary samples

### Biobank: blood sampling, handling and storage

Upon ICU admission, biobank blood samples were collected from patients using Ethylenediaminetetraacetic acid (EDTA) treated test tubes. The samples were then centrifuged in the local hospital laboratory, aliquoted, and subsequently frozen at −80°C. If there was a delay in the freezing process, the samples were kept refrigerated to maintain integrity. Ultimately, the blood samples were stored at the Swecrit biobank in Region Skåne, Lund, Sweden.

In line with the ethical approval (EPN 2015/267 and 2017/802, Lunds University, Lund, Sweden), samples were drawn, collected, and stored without patient consent. An opt-out procedure was used, in which patients discharged from the hospital received a letter 2-3 months after ICU care with information about the study. All patients could choose not to participate by contacting a study nurse or physician. Study data were deleted, and blood samples were destroyed if patients opposed participation in the study.

Patients were excluded from the study for the following reasons: 1) samples were either not collected or were incorrectly labelled, 2) patients were transferred between participating ICU s without new blood samples being drawn, and 3) the patient decided to withdraw from the study.

Regarding NGAL kinetics, additional samples were excluded based on specific sample handling criteria as outlined by Pedersen et al. (2010) [38]:

- Samples stored at room temperature for more than 72 hours post-collection
- Samples stored in a refrigerator for more than 7 days post-collection
- Samples exhibiting hemolysis

After the collection period, all samples were sent to the Department of Clinical Chemistry, Uppsala University Hospital, Uppsala, Sweden, where they were thawed and analysed.

Enzyme-linked immunosorbent assay (ELISA) analyses of NGAL were carried out using a commercial sandwich immunoassay kit (DY1757, R&D Systems, Minneapolis, MN, USA). A monoclonal antibody specific for human NGAL was coated onto microtiter plates. Samples and standards were pipetted into the wells and incubated for 2 h at room temperature to allow NGAL to bind to the immobilised antibodies. After washing, a biotinylated NGAL-specific antibody was added and incubated for 2 h at room temperature. Following a further washing step, a streptavidin–HRP conjugate was added, and the plates were incubated for 30 min. After a final washing step, substrate solution was added, and the enzymatic reaction was stopped by lowering the pH. Absorbance was measured using a SpectraMax 250 microplate reader (Molecular Devices, Sunnyvale, CA, USA).

Concentrations were determined by comparing the optical density of the samples with the standard curve from the same plate. All assays were calibrated against highly purified recombinant human NGAL. The assay primarily detects the monomeric and dimeric forms of NGAL and exhibits no cross-reactivity with MMP-9. All measurements were performed in a blinded fashion, without knowledge of clinical data.

The measuring range of the NGAL assay was 78.1–5,000 pg/mL. Samples were diluted with 1% bovine serum albumin in 0.02 M Na_2_PO_4_, 0.15 M NaCl, pH 7.2, to achieve concentrations within the assay’s measuring range. The basic dilution was 1:50; samples outside this range were re-analysed at adjusted dilutions. The intra-assay variation was 4% and the total variation 6%. We avoid the term “inter-assay variation” as it is sometimes used inconsistently to denote either the total coefficient of variation or only the between-assay component. We therefore use the terms “intra-assay variation” and “total variation”, following the relation: intra-assay variation^2^ + inter-assay variation^2^ = total coefficient of variation^2^.

CRP was analysed using a Particle Enhanced Turbidimetric Assay (PETIA) with reagents from Abbott Laboratories (reagent 6K26-41 and calibrator 6K26-10; Abbott Park, IL, USA) on a Mindray BS380/BS430 chemistry analyser (Mindray, Shenzhen, China). This high-sensitivity assay can detect CRP levels as low as 0.5 mg/L. The following settings were used: R1 90 µL, R2 60 µL, sample 4 µL, positive kinetic reaction type, reaction time positions 24–34. Results were reported with two decimal places. The total coefficient of variation (CV) was 1% (within-run CV 0.8%) at 20 mg/L and 1% (within-run CV 0.6%) at 73 mg/L.

All biomarker analyses were performed by investigators blinded to the clinical data.

In contrast to NGAL, routine CRP values were available to the clinicians.

### Statistical methods

All statistical analyses were performed using R version 4.4.2 [39]. P-values <0.05 were considered statistically significant.

Median values and Interquartile Range (IQR) were reported for continuous variables. The mean value and Standard Deviation (SD) were reported for SOFA scores. The The Mann-Whitney U test was used to assess the difference between non-sepsis and sepsis in independent continuous variables. Differences in proportions were evaluated using Pearson’s *χ*^2^ test.

Associations between NGAL, CRP, and sepsis were analysed using generalised linear models. The discriminatory ability of the biomarkers to distinguish sepsis from non-sepsis was assessed by Receiver Operating Characteristics (ROC) curve analysis, using the pROC package [40]. A bootstrap test with 1,000 replicates was used to compare AUC values.

A Directed Acyclic Graph (DAG) was constructed to map presumed causal relations between sepsis, AKI, CKD, and plasma NGAL, see Fig 1. The DAG includes potential noninfectious causes of AKI that may also affect NGAL. Based on the DAG, we adjusted primary models for baseline CKD (a potential confounder). Still, we did not adjust for AKI or concurrent eGFR because these variables lie on the causal pathway from sepsis to NGAL, and adjustment would constitute overadjustment.

**Fig 1.**
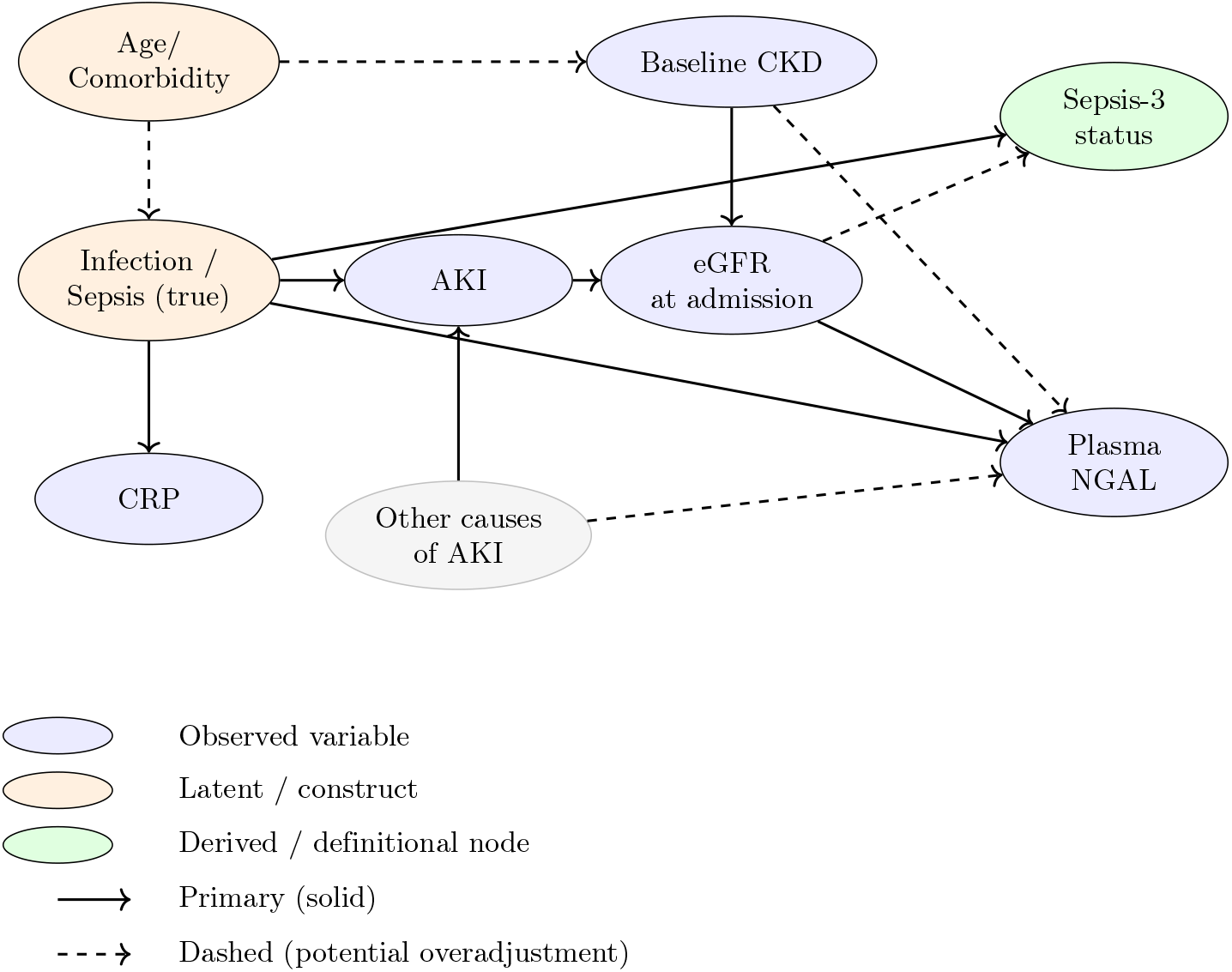
Directed acyclic graph (DAG) illustrating assumed causal relationships between infection, kidney function, NGAL, and sepsis classification.

We report stratified results by kidney function subgroup (No kidney injury; AKI only; CKD only; CKD and AKI) to describe diagnostic performance across clinical phenotypes, noting that stratification conditions on AKI may induce selection/collider bias.

Likelihood ratio tests were conducted to evaluate the interaction between CKD and age.

NGAL was analysed as a continuous variable without applying a predefined test positivity threshold.

For the regression analyses, NGAL was log10-transformed and z-normalised, and CRP was fourth-root transformed and z-normalised to address data skewness. Age was also z-normalised. These transformations were applied to mitigate data skewness and facilitate a more straightforward comparison of results.

Spearman’s rank correlation was applied for correlation analyses.

Sensitivity analysis of NGAL as a diagnostic biomarker for sepsis was performed by stratifying the study population based on kidney function status. Patients were categorised as follows: (1) CKD with AKI, patients with pre-existing CKD who presented with an acute worsening of kidney function at ICU admission; (2) CKD only, patients with CKD but no acute deterioration at admission; (3) AKI only, patients with a normal pre-admission eGF R who developed AKI at ICU admission; and (4) no kidney injury, patients without CKD and without AKI.

To illustrate the correlation between NGAL, CRP and sepsis, Locally Estimated Scatterplot Smoothing (LOESS) regression was calculated using the ggplot2 package [41].

### Bias

CRP levels were available to clinicians in the ICU, which could contribute to the diagnosis of sepsis. This might introduce a bias, resulting in a higher AUC for CRP as a diagnostic marker for sepsis. To reduce confirmation bias in evaluating the diagnostic capability of CRP, we implemented infection criteria that include all culture-positive patients. For culture-negative sepsis, we used blood culture sampling and antibiotic administration as proxy criteria, combined with assessments according to the LMCI.

Our research group previously conducted a study on the same population, with a dropout analysis performed to determine whether missing biobank samples occurred at random.

This study exclusively includes critically ill patients from general intensive care units.

### Missing data

No data for plasma NGAL, CRP, creatinine, age, or sex were missing. Creatinine measurement obtained 7–365 days before ICU admission was missing in 38% of the study population.

## Results

### Participants

During the study period, a total of 8360 ICU patients were assessed, of which 4732 met the inclusion criteria. Among these, 2071 patients (44%) satisfied the criteria for sepsis. Notably, 81% of the sepsis cases were identified based on positive culture results, while the remaining 19% met clinical criteria for infection despite negative cultures. For a detailed overview, see Fig 2.

**Fig 2.**
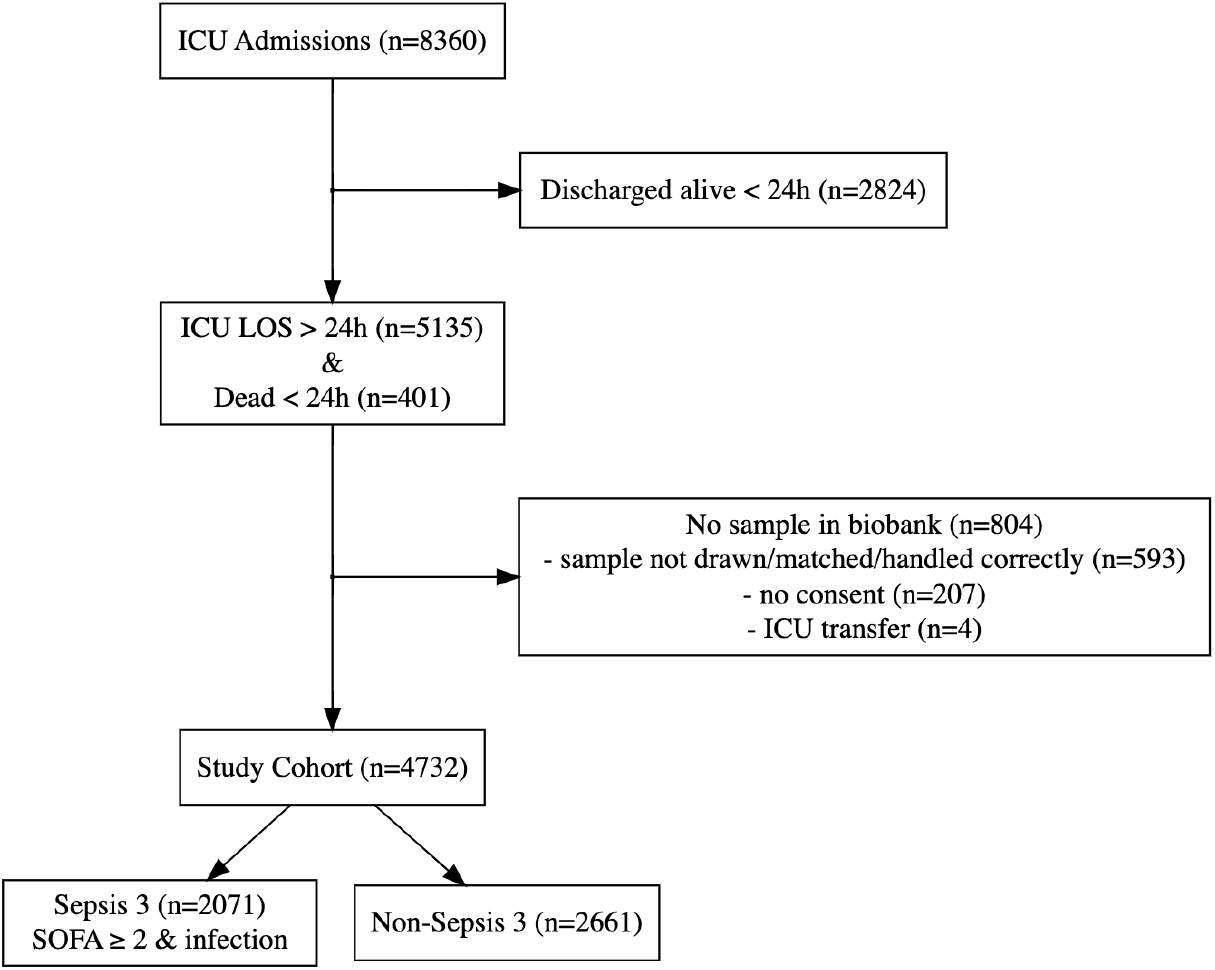
Flow chart of patient inclusion and exclusion. The included cohort had an ICU length of stay of >24 hours or died *≤*24 hours after ICU admission and had eligible biobank blood samples. Of these, 44% had sepsis. Infection criteria were culture-positive *±* 48 hours or blood culture & antibiotics & probable infection according to LMCI. ICU, intensive care unit; LMCI, Linder–Mellhammar criteria of infection.

### Descriptive data

Patients with sepsis exhibited a higher total SOFA score (mean 8.3 vs. 6.3, p <0.001), compared to non-sepsis patients, reflecting a greater degree of organ failure. Patients with sepsis also had a longer ICU length of stay (median 3.3 vs. 2.0 days, p <0.001), and higher 30-day mortality (33% vs. 28%, p <0.001). A reduced eGFR was more common in patients with sepsis (61% vs. 47%, p <0.001), as was the use of Continuous Renal Replacement Therapy (CRRT) (18% vs. 9.5%, p <0.001), and the frequency of invasive ventilation (69% vs. 60%, p <0.001).

Plasma NGAL levels were significantly higher in patients with sepsis compared to those without (median 315 ng/mL, IQR 156–660 ng/mL vs. 170 ng/mL, IQR 101–309 ng/mL), as were CRP levels (median 95 mg/L, IQR 26–186 mg/L vs. 17 mg/L, IQR 3–69 mg/L), see Table 1.

**Table 1.** Descriptive characteristics and clinical outcomes of patients with and without sepsis.

|  | Non-sepsis | Sepsis | P-value |
| --- | --- | --- | --- |
| **n** | 2661 | 2071 | – |
| Age, years (median [IQR]) | 67 [54–75] | 69 [59–76] | <0.001 |
| Male sex (%) | 1621 (61) | 1235 (60) | 0.39 |
| **Before ICU admission** |  |  |  |
| ICU admission <24 h from hosp. adm. (%) | 1613 (61) | 1220 (59) | 0.25 |
| ICU admission >48 h from hosp. adm. (%) | 658 (25) | 538 (26) | 0.34 |
| Admitted from ER (%) | 1121 (42) | 835 (40) | 0.22 |
| Admitted from hospital ward (%) | 710 (27) | 848 (41) | <0.001 |
| Admitted from OR (%) | 527 (20) | 173 (8.4) | <0.001 |
| **Biomarkers at ICU admission** |  |  |  |
| NGAL, ng/mL (median [IQR]) | 170 [101–309] | 315 [156–660] | <0.001 |
| C-reactive protein, mg/L (median [IQR]) | 17 [3–69] | 95 [26–186] | <0.001 |
| Lactate, mmol/L (median [IQR]) | 2.0 [1.1–3.9] | 2.2 [1.2–4.3] | <0.001 |
| WBC, 10<sup>9</sup>/L (median [IQR]) | 12.7 [9.0–17.3] | 13.2 [8.2–19.2] | 0.19 |
| Leukopenia (%) | 116 (4.4) | 266 (13) | <0.001 |
| Neutropenia (%) | 15 (0.6) | 68 (3.3) | <0.001 |
| **Physiology at ICU admission** |  |  |  |
| Shock (%) | 747 (28) | 773 (37) | <0.001 |
| Body temperature, °C (median [IQR]) | 37.0 [36.0–37.4] | 37.0 [36.3–38.0] | <0.001 |
| Body temperature >38 °C (%) | 233 (9) | 439 (21) | <0.001 |
| Hypothermia (%) | 475 (18) | 331 (16) | 0.08 |
| eGFR <60 mL/min/1.73 m<sup>2</sup> (%) | 1250 (47) | 1273 (61) | <0.001 |
| **Outcomes** |  |  |  |
| ICU LOS, days (median [IQR]) | 2.0 [1.3–3.8] | 3.3 [1.8–6.5] | <0.001 |
| ICU mortality (%) | 468 (18) | 403 (20) | 0.11 |
| 30-day mortality (%) | 749 (28) | 686 (33) | <0.001 |
| CRRT (%) | 252 (9.5) | 375 (18) | <0.001 |
| Invasive ventilation (%) | 1608 (60) | 1421 (69) | <0.001 |

ICU, intensive care unit; LOS, length of stay; ER, emergency room; OR, operating room; IQR, interquartile range; NGAL, neutrophil gelatinase-associated lipocalin; CRP, C-reactive protein; WBC, white blood cell count; SOFA, Sequential Organ Failure Assessment; CRRT, continuous renal replacement therapy.

As shown in (Fig S1), higher NGAL and CRP concentrations were generally associated with a higher probability of sepsis.

NGAL and CRP were positively correlated (*ρ* = 0.403, p <0.001), see (Fig S2).

### Unadjusted and adjusted linear models

A Generalised Linear Model (GLM) evaluating NGAL as a diagnostic marker of sepsis yielded an Odds Ratio (OR) of 1.90 (95% CI): 1.78–2.03, p <0.001) and a ROC AUC of 0.67 (95% CI: 0.66–0.69).

CRP demonstrated superior discriminatory ability for sepsis compared to NGAL, with an AUC of 0.72 (95% CI: 0.71–0.73, p <0.001). The combination of NGAL and CRP, analysed using GLM and ROC analysis, showed improved diagnostic performance compared to CRP alone, with an AUC of 0.74 versus 0.72 (p <0.001), (Fig 3 and Fig 4).

**Fig 3.**
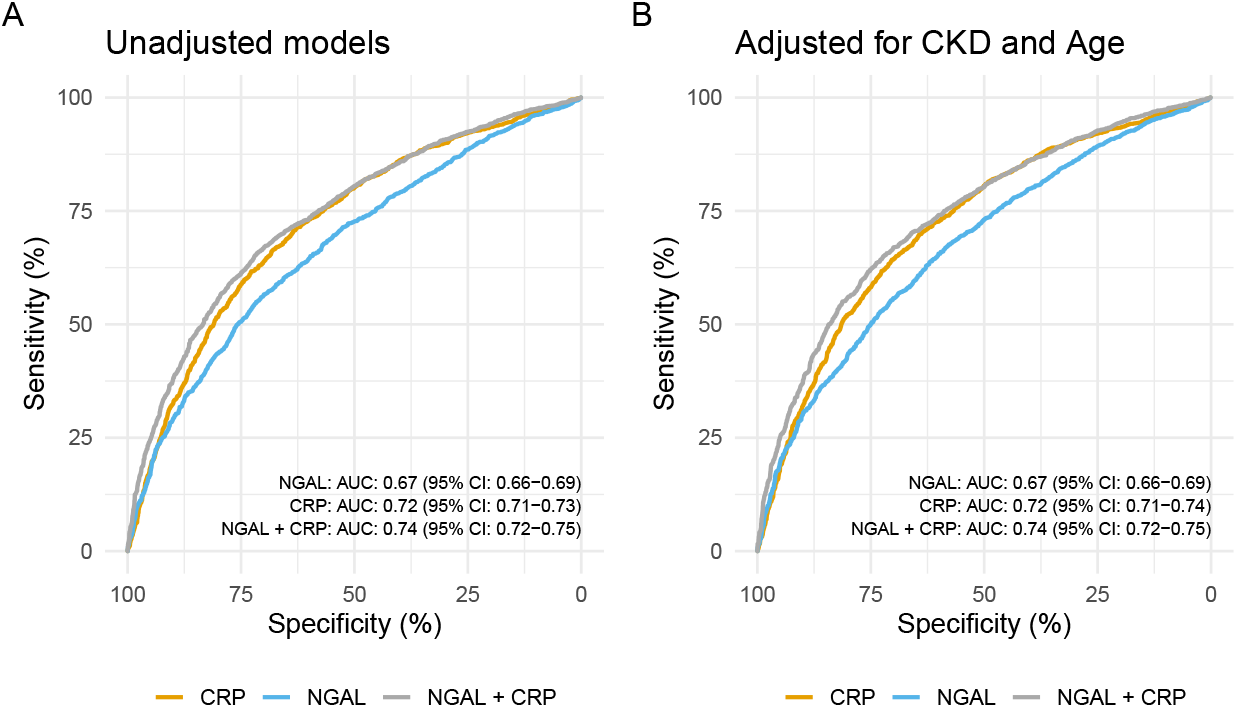
Receiver operating characteristic analysis for NGAL and CRP. (A) Receiver operating characteristic (ROC) curves and corresponding areas under the curve (AUCs) with 95% CIs for NGAL, CRP, and their combination as predictors of sepsis. (B) ROC AUCs with 95% CIs for NGAL, CRP, and their combination adjusted for chronic kidney disease and age. NGAL, neutrophil gelatinase–associated lipocalin; CRP, C-reactive protein; AUC, area under the curve; CI, confidence interval; CKD, chronic kidney disease.

**Fig 4.**
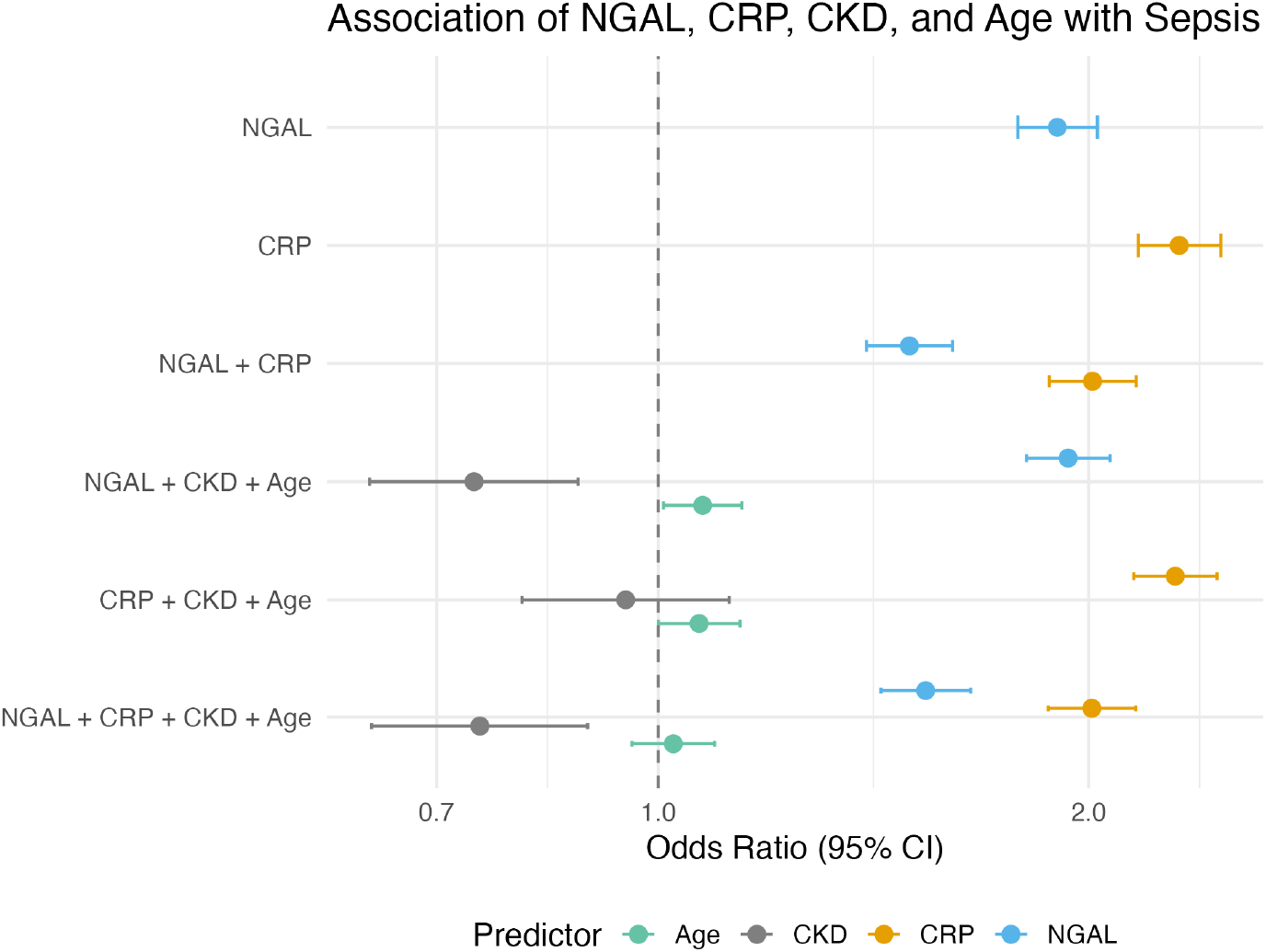
Odds ratios for sepsis associated with NGAL and CRP. Forest plot showing odds ratios (ORs) with 95% CIs for NGAL and CRP as predictors of sepsis based on generalised linear models. The models include: (1) NGAL alone; (2) CRP alone; (3) NGAL and CRP; (4) NGAL adjusted for chronic kidney disease and age; (5) CRP adjusted for chronic kidney disease and age; and (6) NGAL and CRP adjusted for chronic kidney disease and age. NGAL, neutrophil gelatinase-associated lipocalin; CRP, C-reactive protein; OR, odds ratio; CI, confidence interval; CKD, chronic kidney disease.

The discriminative ability of NGAL for sepsis remained essentially unchanged after adjustment for CKD and age. Similarly, combining NGAL with CRP yielded comparable diagnostic performance regardless of these adjustments.

Likelihood ratio tests (LRT) confirmed that including CKD and age as interaction terms did not significantly improve model fit compared to using them as adjustment variables (CKD p = 0.85, age p = 0.12).

### Sensitivity analyses

We evaluated whether adding NGAL to CRP improved sepsis discrimination within subgroups stratified by kidney function status. As shown in Fig 5, the discriminatory performance of NGAL and CRP varied across kidney function subgroups. The combination of NGAL and CRP demonstrated superior diagnostic performance compared with CRP alone in patients without kidney injury (p = 0.001) and in those with AKI only (p <0.001). Among patients with CKD only and those with both CKD and AKI, the differences in AUC did not reach statistical significance (p = 0.052 and p = 0.081, respectively), (Fig 5).

**Fig 5.**
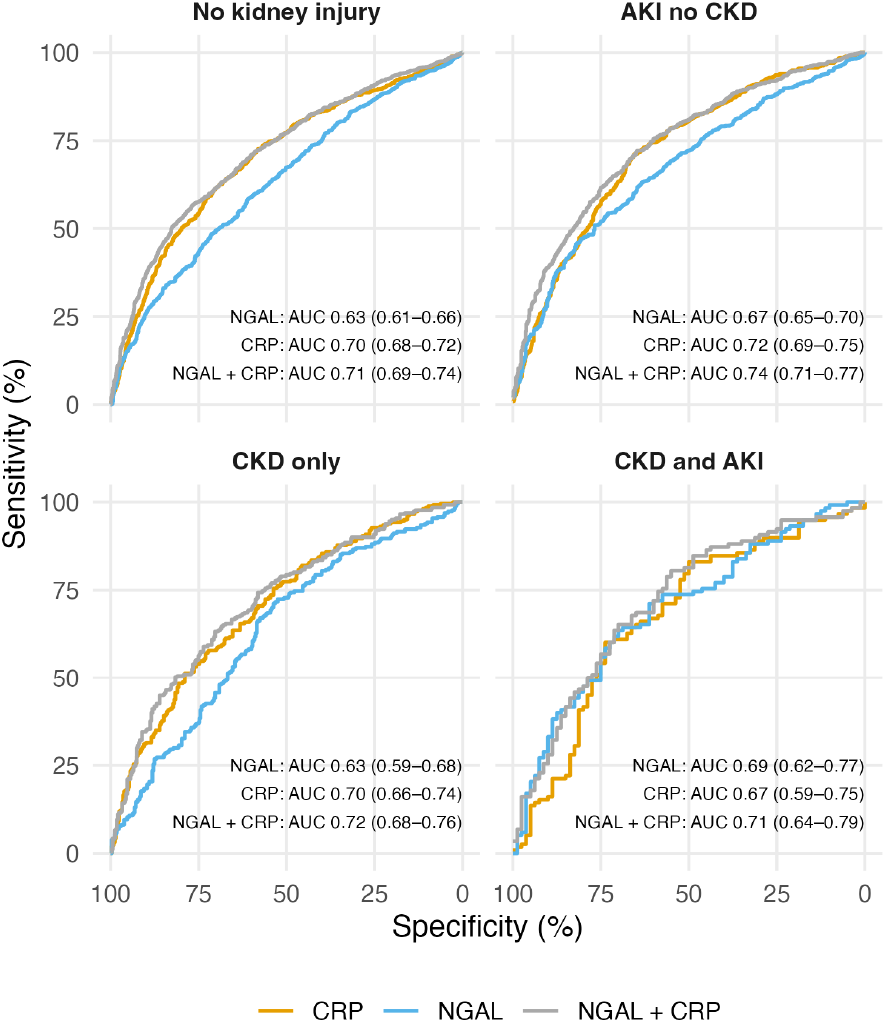
Receiver operating characteristic analysis stratified by kidney function. Receiver operating characteristic (ROC) curves and corresponding areas under the curve (AUCs) with 95% CIs for NGAL, CRP, and their combination for sepsis prediction across kidney function subgroups. NGAL, neutrophil gelatinase-associated lipocalin; CRP, C-reactive protein; AUC, area under the curve; CI, confidence interval.

### Dropout analysis

Our research group previously investigated this population and conducted a dropout analysis to assess whether missing biobank samples were random. This analysis showed that 5536 patients had an ICU length of stay greater than 24 hours or died within 24 hours. Out of these patients, 804 (15%) lacked biobank blood samples, resulting in their exclusion from the study population. Compared to the excluded patients, the study population had a higher degree of sepsis (44% vs. 32%, p <0.001) and shock (32% vs. 27%, p=0.004), and presented with greater overall illness severity (SOFA score 7.2 vs. 6.4, p <0.001), yet demonstrated a lower 30-day mortality rate (30% vs 35%, p=0.008). A higher proportion of the included patients were admitted directly to the ICU from the emergency room (41% vs. 37%, p=0.012). Among routinely collected biomarkers at ICU admission, lactate levels were higher (2.0 vs 1.8 mmol/L, p=0.031), whereas CRP levels were consistent (43 vs 42 mg/L, p=0.28).

## Discussion

This multicenter study assessed NGAL as a diagnostic marker for sepsis in a large cohort of critically ill patients. We found that NGAL demonstrated moderate discriminatory power, with an ROC AUC of 0.67. CRP outperformed NGAL (AUC 0.72 vs. 0.67, p <0.001). However, the combination of NGAL and CRP significantly enhanced diagnostic accuracy compared to CRP alone (AUC 0.74 vs. 0.72, p <0.001), indicating that NGAL provides additional information beyond CRP.

CKD and age were treated as confounders because they may affect both baseline NGAL levels and susceptibility to infection. In contrast, AKI may lie on the causal pathway from infection to NGAL elevation, and adjusting for AKI could obscure the total diagnostic signal of infection and introduce overadjustment or collider bias. Consequently, by limiting adjustment to CKD and age, we aimed to capture the full diagnostic contribution of NGAL.

Adjustment for CKD and age did not alter the diagnostic performance of NGAL, indicating that NGAL provides sepsis-related information largely independent of these covariates.

Research on NGAL as a diagnostic marker for sepsis is limited [19, 21–25]. Several studies focus on NGAL as a predictor of sepsis-associated AKI, though its efficacy has been debated [15]. Notably, the 28th Acute Disease Quality Initiative workgroup (2023) identifies NGAL as an accurate predictor of sepsis-associated AKI [16].

Sepsis and AKI are closely linked, with about 70% of sepsis patients developing AKI [10]. Sepsis is the leading cause of AKI among hospitalised patients [42]. In this context, a biomarker like NGAL, which acts as an acute-phase protein involved in the innate immune system and serves as a rapid indicator of kidney function, is particularly appealing for sepsis diagnosis. On the other hand, kidney injury may arise from causes unrelated to sepsis, which can complicate the interpretation of NGAL levels specifically as a sepsis biomarker.

Today, serum creatinine is included in sepsis diagnostics, as it is incorporated into the SOFA score. However, creatinine has several limitations. Firstly, acute changes in GFR are not immediately reflected in serum creatinine levels, as it takes days for the balance between creatinine production and elimination to stabilise. Consequently, serum creatinine often underestimates the degree of renal function loss, especially within the first 48 hours after an insult [12, 43]. Secondly, creatinine production is reduced during sepsis [44].

The search for a highly specific sepsis diagnostic marker is challenging due to the heterogeneity of sepsis. Efforts are underway to identify subgroups within the sepsis population to improve diagnostic accuracy. Bhavani et al. recently proposed that subgrouping sepsis cases by temperature trajectories reveals significant differences in mortality and biomarker levels among these subphenotypes, with NGAL levels peaking in the hypothermic group [45].

Our results are more modest than previous studies showing NGAL’s potential as a sepsis marker in critically ill patients. For example, Mårtensson et al. found NGAL to diagnose sepsis regardless of kidney function, with an AUC of 0.82 for bacterial infection detection [19]. Md Ralib et al. reported an AUC of 0.71 for differentiating sepsis from noninfectious Systemic Inflammatory Response Syndrome (SIRS) [21]. A recent article by Abdelrazic et al. demonstrated that elevated serum NGAL levels at admission were linked to increased sepsis severity and mortality in critically ill children in the pediatric ICU. The study showed that NGAL outperformed the pediatric risk of mortality III (PRISM III) score as a predictor of outcomes in bacterial sepsis [22].

We found that NGAL provided a statistically detectable improvement in diagnostic accuracy compared with CRP alone; however, the magnitude of this improvement was minor. The cohort size strengthens the precision of our estimates, and the minimal improvement observed indicates that NGAL is unlikely to play a significant role in routine sepsis assessment in the ICU.

### Strengths and limitations

A key strength of this study is its large, multicentre patient cohort, which enhances the generalisability of the findings across diverse ICU settings. Furthermore, the medical records of all culture-negative patients with suspected infection were meticulously reviewed for clinical and laboratory evidence of infection using the LMCI criteria. This enabled a more accurate distinction between true sepsis and non-sepsis cases, reducing potential misclassification bias. Another methodological strength is the retrospective classification of infection based on microbiological cultures obtained around the time of NGAL measurement. Because culture results are typically delayed, this approach evaluates NGAL against a reference standard that is temporally concurrent but not yet known at the time of testing, thereby closely reflecting its intended clinical use for early sepsis detection.

Because AKI has multiple causes (including both sepsis and noninfectious factors), AKI functions as a collider in the DAG (Sepsis → AKI ← Other causes). Conditioning on AKI (e.g., by stratification or adjustment) may open non-causal paths and induce bias; results within AKI strata should be interpreted with caution.

A limitation of this study is the measurement of NGAL at a single time point after ICU admission, without serial sampling to capture temporal trends in the ICU. Previous work by Jonsson et al. has demonstrated that NGAL levels decrease following the initiation of antibiotic therapy [46], suggesting that dynamic assessment might provide additional prognostic or diagnostic value.

Finally, as the present study includes only critically ill patients admitted to general intensive care units, the findings may not be fully generalisable to patients with milder sepsis or early infections managed outside the ICU environment.

## Conclusion

In this large cohort of critically ill patients, NGAL showed modest discrimination for sepsis. Combining NGAL with CRP offered a small improvement in model performance, but the clinical impact appears limited. Our findings do not support using NGAL for sepsis diagnosis in the ICU.

## Declarations

### Ethics approval and consent to participate

The Regional Ethical Review Board of Lund approved the study protocol, Sweden (registration no. 2017/802 and 2015/267), before the study was conducted. Following the ethics approval, consent for blood sampling to the biobank was presumed. After 2-3 months, survivors received a letter with information about the opportunity to opt out. The study was conducted in accordance with the tenets of the Declaration of Helsinki.

## Consent for publication

Not applicable.

## Availability of data and materials

Public access to the data underlying the results presented in this study is restricted under the Swedish Public Access to Information and Secrecy Act due to the inclusion of potentially identifying and sensitive patient information. Data may be made available for research purposes following approval by the Swedish Ethical Review Authority and the relevant data-holding authorities in Region Skåne, Sweden.

## Authors’ contributions

Attila Frigyesi and Maria Lengquist conceived the study. Attila Frigyesi and Hans Friberg funded the study. Anders Olof Larsson analysed NGAL. Attila Frigyesi, Maria Lengquist, and Lisa Boström designed the study. Lisa Boström and Sofie Hagström performed all the calculations and produced all the figures and tables. Lisa Boström and Sofie Hagström wrote the first draft of the manuscript. Lisa Boström, Sofie Hagström, Jonas Engström, Anders Olof Larsson, Hans Friberg, Maria Lengquist and Attila Frigyesi contributed to the final manuscript.

## Supporting information

**Fig S1.**
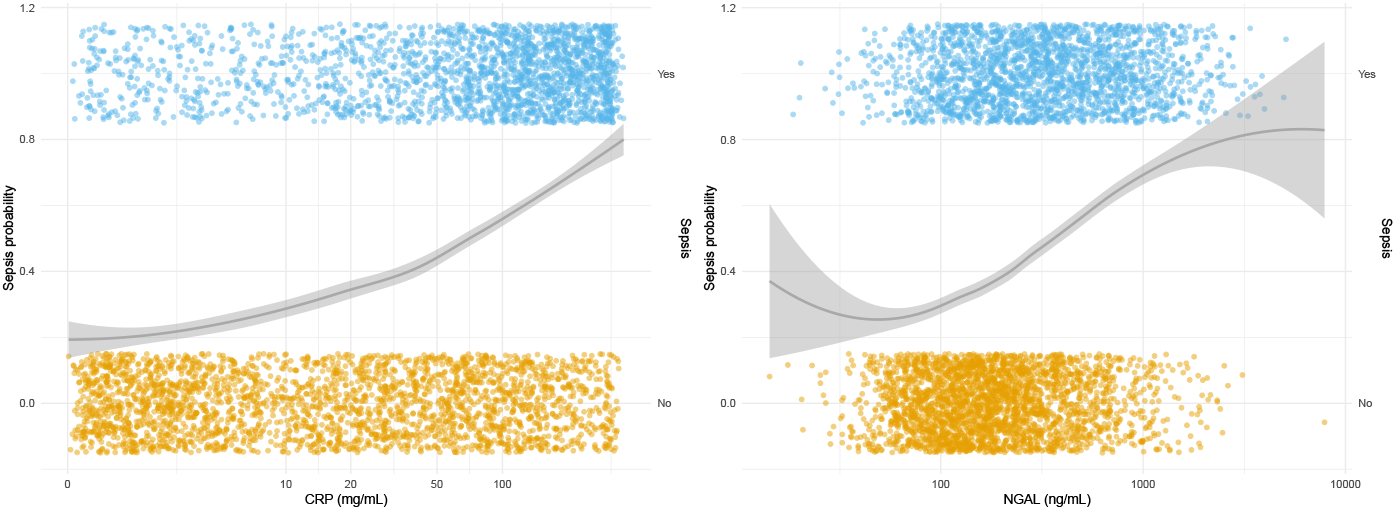
The probability of sepsis in relation to NGAL and CRP. The solid line (LOESS) shows the probability of sepsis (left y-axis) in relation to NGAL and CRP. Points indicate sepsis or no sepsis (right y-axis). NGAL, neutrophil gelatinase–associated lipocalin; CRP, C-reactive protein.

**Fig S2.**
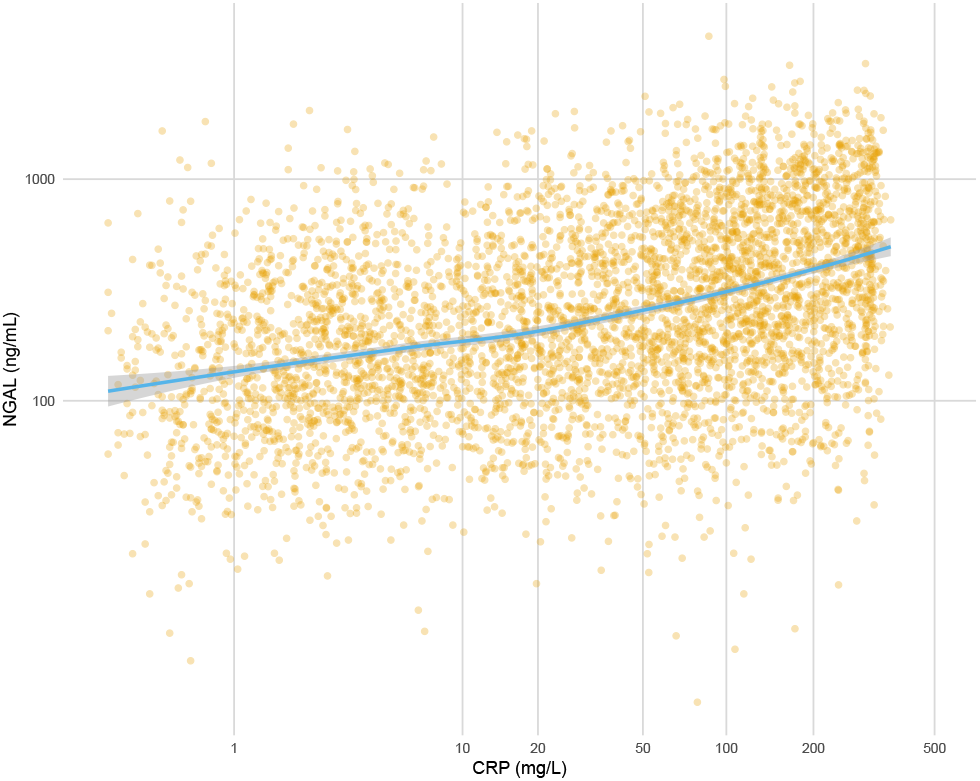
Relationship between NGAL and CRP. Scatterplot of NGAL and CRP, with NGAL on log10 scale and CRP on fourth-root scale. Solid line is local polynomial regression fit. NGAL, neutrophil gelatinase–associated lipocalin; CRP, C-reactive protein.

## Acknowledgments

We thank all staff at the ICUs of Skåne University Hospital in Malmö and Lund, Helsingborg Hospital, and Kristianstad Hospital for contributing to this study.

